# Study of Microbial Profile, Sensitivity Pattern of Community-Acquired Pneumonia in Diabetic Patients and Correlation with Glycaemic Status

**DOI:** 10.1101/2025.03.20.25324334

**Authors:** Md Mahmudunnaby, Mahmudul Hasan Nahid, Mustari Sarkar Trisha

## Abstract

**Background:** Pneumonia, an infection of the lung parenchyma, is classified into community-acquired pneumonia (CAP) and nosocomial pneumonia. CAP occurs in patients who have not been hospitalized or resided in long-term care facilities for more than 14 days before symptom onset. Diabetes mellitus (DM), an immunosuppressive condition, predisposes patients to severe infections, including pneumonia. Diabetic patients with CAP often encounter more virulent or atypical pathogens and show increased resistance to conventional antibiotics. This study aimed to identify the bacterial etiology of CAP in diabetic patients and assess their antibiotic sensitivity patterns.

**Methods:** A descriptive cross-sectional study was conducted in the Department of Medicine, Shaheed Suhrawardy Medical College Hospital, Dhaka, from 29th December 2017 to 28th June 2018. A total of 100 diabetic patients with CAP were enrolled following a structured protocol that included history-taking, clinical examination, and relevant investigations. Sputum samples were analyzed using standard microbiological techniques. Data were processed and analyzed using SPSS version 22 and Microsoft Excel. Quantitative data were expressed as mean ± standard deviation, while qualitative data were presented as frequency and percentage.

**Results:** The majority of the patients (54%) were between 51–60 years, with a mean age of 49.7 ± 9.23 years. Of the 100 cases, 62% were male, and 38% were female, with a male-to-female ratio of 1.6:1. High-grade fever, sweating, and cough were the most common symptoms. Sputum cultures revealed growth in 92% of samples. The predominant pathogens were *Klebsiella pneumoniae* (30.4%), *Streptococcus pneumoniae* (23.9%), and *Staphylococcus aureus* (13.0%). Antibiotic resistance was highest against ceftriaxone and amoxicillin, with variable sensitivity across isolates.

**Conclusion:** The study concludes that poor glycemic control is associated with increased susceptibility to infections. Early detection, appropriate management, and strict glycemic control can reduce the burden of pneumonia in diabetic patients.

**Lay Summary:** *Understanding the Relationship Between Pneumonia and Diabetes:* Pneumonia is a serious lung infection causing coughing, fever, and breathing difficulties. People with diabetes are more vulnerable to infections due to weakened immune systems. This study, conducted in Bangladesh, explores how diabetes affects the bacteria causing pneumonia and their resistance to antibiotics.

**Key Findings:**

- The most common bacteria in diabetic pneumonia patients were *Klebsiella pneumoniae* (30.4%), *Streptococcus pneumoniae* (23.9%), and *Staphylococcus aureus* (13.0%).
- Antibiotics that lost efficacy due to resistance were highest for ceftriaxone and amoxicillin.
- Poor blood sugar control led to more infections and antibiotic resistance.

**Why It Matters:** These findings highlight the need for strict blood sugar control and careful antibiotic selection to reduce pneumonia risks in diabetic patients.

## Introduction

Community-acquired pneumonia (CAP) is the leading cause of death among infectious diseases and an important health problem, having considerable implications for healthcare systems worldwide. Typical bacterial pathogens that cause CAP include *Streptococcus pneumoniae*, *Haemophilus influenzae*, and *Moraxella catarrhalis*. CAP management still has significant drawbacks. Mortality remains very high in severely ill patients presenting with respiratory failure or shock but is also high in the elderly and patients having immunosuppressive condition, diabetes mellitus and presence of comorbidity.

Diabetes mellitus is an immunosuppressive state leading to increased susceptibility to various infections. Pneumonia, urinary tract infections, skin & soft tissue infections are all more common in the diabetic population. Globally, an estimated 422 million adults were living with diabetes, compared to 108 million in 1980. The global prevalence (age-standardized) of diabetes has nearly doubled since 1980, rising from 4.7% to 8.5% in the adult population^1^. The global costs of diabetes and its consequences are large and will substantially increase by 2030^2^. A positive association presents diabetes and infection. Bacterial infections are a relatively frequent occurrence in diabetic patients and there may be an associated increase in morbidity and mortality.

Worldwide the estimated prevalence of diabetes was 4.0% in 1995 and to rise to 5.4% by the year 2025. The number of adults with diabetes in the world will rise from 135 million in 1995 to 300 million in the year 2025^3^. The high incidence of Diabetes in Bangladesh is due to a sedentary lifestyle, lack of physical activity, obesity, stress, and consumption of a diet rich in fat, sugar, and calories. Increasing economic growth will raise Diabetes prevalence in Bangladesh even more than what is estimated. Data suggest that Indians as an ethnic group have a high risk of type II DM most likely due to genetic susceptibility. So, diabetes-specific complications are also more common in developing countries than in other races^4, 5^. Pneumonia, urinary tract infection, and skin & soft tissue infection are all more common in the diabetic population^4^.

In general, the organisms that cause pulmonary infection are like those found in the non-diabetic population; however, gram-negative organisms, *Staphylococcus aureus* and *Mycobacterium tuberculosis* are more frequent organisms^6^. For patients with community-acquired pneumonia, diabetes mellitus is one of the most common predisposing risk factors. The magnitude and duration of hyperglycemia with glycaemic status are strongly associated with the severity of microvascular and neurologic complications^7^.

The presence of hyperglycemia increases the risk of infection. The predisposition for infection may also be based on conditions that interfere with normal clearance mechanisms and on disturbance of pulmonary immune cell function^5^. In patients with pneumonia, Diabetes Mellitus is associated with polymicrobial etiology, multilobe involvement, increased ICU admissions, and increased severity in the form of high PSI score and mortality^8^. Glycemic status has an impact on the severity of complications and infection. Specific defects in innate and adaptive immune function have been identified in diabetic patients in a range of in vitro studies. However, the relevance of these findings to the integrated response to infection in vivo remains unclear, especially in patients with good glycaemic control. Vaccine efficacy seems adequate in most diabetic patients, but those with type 1 diabetes and high glycosylated hemoglobin levels are most likely to exhibit hypo responsiveness. While infections are closely associated with diabetes, this is usually in the context of extreme metabolic disturbances such as ketoacidosis. The link between glycaemic control and the risk of common community-acquired infections is less well established but could be clarified if infection data from large community-based observational or intervention studies were available^9^.

Several aspects of immunity such as polymorphonuclear leukocyte function (i.e. leukocyte adherence, chemotaxis, and phagocytosis) and bactericidal activity of serum are depressed in patients with diabetes^10^. Alteration in T-lymphocyte subsets, including a relative reduction in T-helper lymphocytes, could interfere with immune defense against infection. As a response to infection and cytokine release, insulin resistance in peripheral tissue occurs, resulting in the elevation at blood Sugar^7^. Hyperglycemia impairs a wide range of functions in neutrophils & monocytes (macrophages). This is particularly important in limiting invasion by pyogenic and other bacteria^7^. Adherence and phagocytosis depend on the recognition of specific molecules on the bacterial surface including bacterial glycoproteins as well as attached complement and IgG produced as a result of the immune response to the infection. The movement of phagocytic cells to the sites of infection is generally impaired in diabetes but improves with glycemic control^11^.

The presence of healthy microcirculation is essential to certain infectious insults. Alteration in the function of capillary endothelium, the rigidity of red blood corpuscles and changes in the oxygen dissociation curve that occur as a result of chronic hyperglycemia are factors that affect the host’s ability to combat infection. It is therefore no surprise that patients with long-standing diabetes with microvascular complications are at a much greater risk of infections than non-diabetic or diabetics without complications. The reduced oxygen supply to tissue as a result of microvascular changes predisposes them to infections by anaerobic organisms which grow best under such conditions^11^.

A study in Bangladesh reported that most of the patients had growth of *Klebsiella pneumoniae* in sputum, followed by *Staphylococcus aureus* and then other gram-negative bacteria. All (100%) of the *Pseudomonas* and *Acinetobacter* were sensitive to colistin, and all (100%) of the Staphylococcus aureus were sensitive to vancomycin. Regarding glycemic status, most of the bacterial growth was isolated in patients with uncontrolled DM as evidenced by HbA1c ≥7.0%^12^. This is because uncontrolled DM causes immunosuppression leading to an increased chance of any infection including pneumonia. Therefore, the study aimed to see the microorganisms most commonly causing community-acquired pneumonia in diabetic patients.

Pneumonia is the major cause of morbidity, mortality, and cost of care worldwide. Because of population growth, aging, and physical inactivity, the prevalence of diabetes is rapidly increasing. Diabetes and hyperglycemia are generally thought to be risk factors for infection. Pneumonia in diabetic patients is often atypical, caused by more virulent organisms, and associated with increased antibiotic resistance. Poor glycemic status or hyperglycemia accelerates the CAP. This is because uncontrolled DM causes immunosuppression leading to an increased chance of any infection including pneumonia. So, the aim of this study is to see sputum culture & drug sensitivity patterns of community-acquired pneumonia in diabetic patients & their correlation with glycogenic status and thus try to find out if there is any association present between glycaemic status and with occurrence of community-acquired pneumonia in diabetes patients.

## Materials and Methods

### 2.1 Study design

A descriptive cross-sectional study.

### 2.2 Place of study

Department of Medicine, Shaheed Suhrawardy Medical College Hospital, Dhaka.

### 2.3 Study periods

29th December 2017 to 28^th^ June 2018.

### 2.4 Study Population

Diabetic patients with community-acquired pneumonia were enrolled for the study.

### 2.5 Sample size

The following standard formula is widely used in determining sample size:

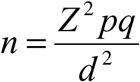

n= the desired sample size

z= Standard normal deviate usually set at 1.96

p= Proportion in the population. In a previous study, the crude incidence of hospitalization for infection was 23.7 (21.3–26.2)/1,000 patient-years. The 368 admissions consisted of pneumonia (nLJ=LJ181, 49.2%), cellulitis (nLJ=LJ107, 29.1%), septicemia/bacteremia (nLJ=LJ42, 11.4%), osteomyelitis (nLJ=LJ19, 5.2%), genitourinary infection (acute pyelonephritis, renal/perinephric abscess or cystitis; nLJ=LJ14, 3.8%) and others (meningococcal disease, otitis media, otitis externa, sinusitis and other bacterial infection; nLJ=LJ5 (1.4%)^31^. So in this study p-value will be 49.2%. or 0.49

q= 1-p, Or 0.51

d= Acceptable error or precision (which is considered as 10%.

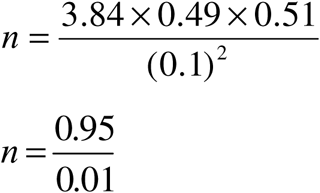

According to this formula, the targeted sample will be 95.9. The current study duration is 6 months; therefore 100 cases will be taken.

### 2.6 Sampling

he sample was selected by a consecutive sampling technique.

### 2.7 Inclusion criteria

- Diabetic patient
- Community-acquired pneumonia

### 2.8 Exclusion criteria

- Nosocomial pneumonia
- Ventilator-associated pneumonia
- Patient on immunosuppressive agent or immunosuppression, defined as chemotherapy or neutropenia <1000/µL during the past 28 days: treatment with more than 20mg corticosteroids daily for more than 14 days, HIV infection, immunosuppressive therapy after organ or bone marrow transplantation.
- Once a diagnosis of Pulmonary Tuberculosis is made, such cases will be excluded from the study of clinical profile.

### 2.9 Operational definition

*Diagnosing CAP*:a. Symptoms of an acute lower respiratory tract illness (cough with or without expectoration, shortness of breath, pleuritic chest pain) for less than 1 week; and b. At least one systemic feature (temperature >37.7°C, chills, and rigors, and/or severe malaise); and c. New focal chest signs on examination (bronchial breath sounds and/or crackles) with. d. No other explanation for the illness.

With new radiographic shadowing for which there is no other explanation (not due to pulmonary edema or infarction). Radiographic shadowing may be seen in the form of a lobar or patchy consolidation, loss of a normal diaphragmatic, cardiac, or mediastinal silhouette, interstitial infiltrates, or bilateral perihilar opacities, with no other obvious cause.

*DM*: The current WHO diagnostic criteria for diabetes

- Fasting blood glucose ≥ 7.0 mmol/l
- Or 2 hours after ingestion of 75 gm glucose ≥ 11.1 mmol/l
- Or HbA1C ≥ 6.5 %

In the presence of classical symptoms of hyperglycemia Or, Diagnosed case of T2DM on anti-diabetic medications

Normal blood glucose level:

- Fasting blood glucose: LJ 6.1 mmol/L
- 2-hour postprandial blood glucose: LJ 7.8 mmol/L
- HbA1C: < 6.0%

Glycemic control: Targets are-

- HbA1c: LJ 7.0%

*Sputum examination*: The procedure of sputum specimen collection is usually non-invasive. Commonly, the "deep cough" sample of the early morning is collected before eating or drinking anything to avoid bias in interpreting the results. Clinical diagnostic sputum tests aim to detect the causes of lower respiratory tract infections and some other diseases. Sputum culture is the most common test that needs to be performed when the patient has pneumonia. It is used to identify bacteria or fungi causing airways or lung infections.

Sputum smear microscopy is the initial step taken in laboratory sputum analysis. The Gram stain is the first staining technique performed in preliminary bacterial identification, which helps determine if there is an adequate number of pathogens in the culture and make a definitive diagnosis.

Sputum Culture Procedure: The sputum sample is added to a culture plate with a specific substance that promotes the growth of bacteria or fungi. Then cover the lid of the dish and place it in a 37-degree C incubator. The lab specialist checks the bacteria or fungi growth in the sputum plate every day. Once the sputum culture is positive, microscopy, colony morphology, or biochemical tests of bacterial growth are performed to identify the specific type of bacterium or fungus.

Sputum Staining Tests Procedure: The sputum specimen is a smear on a microscope slide. Different staining dyes are added to the cells, bacteria, or fungi of the sample on the slides and then washed with water, alcohol, or acid solutions. The slides are then diagnosed under a microscope. If the bacteria, fungi, or specific cells are identified in the specimens, the results are positive.

Sputum Biochemical Tests Procedure: To identify a suspected organism, at first, the bacteria will be inoculated in a series of differential media. Then use different indicators to observe the specific end products of metabolism inside of the medium.

Sputum Cytology Examination Procedure: The smear sputum slide is stained with different dyes according to the instructions. Then the pathology specialist examines the stained slide under the microscope to find the abnormal cells from the sputum specimen.

Sputum Antimicrobial Susceptibility Tests Procedure: For the MIC method, the bacteria or fungi isolated from sputum specimens were diluted in saline and swabbed onto the MIC panels. For the dish diffusion method, selected different concentrations of antibiotics are placed directly onto the bacteria-swabbed agar plates. Panels or plates are incubated at 35 degrees C for about 16 to 18 hours or longer. The minimal concentration of the antibiotic that inhibits the growth of organisms or MIC panel is read according to the guidelines of different manufacturers. Then the result is reported.

*Socioeconomic status*: Socioeconomic status is classified by household income, in accordance with Household Income and Expenditure Survey (HIES)-2010, World Bank report, UNICEF-The State of the World’s Children 2015: Executive Summary, and Statistical Pocketbook of Bangladeh-2013: Bangladesh Bureau of Statistics. Categories are arranged by integration, modifications and accommodation of all reports. The classifications of income group people in Bangladesh are mainly the following types:

- Poor class: Ultra-poor or extremely poor households are under the lower poverty line, as determined by HIES. The income equivalent of the lower poverty line is a maximum monthly income of approximately BDT 5,000 or <5000.
- Non-Poor class: Non-poor households, but often tenuously so, are those whose monthly income is above the upper line. A non-poor household is at or below the nationally declared minimum taxable income. Households having monthly incomes between BDT 5,800 and BDT 19,000 are considered to be in this group, which are also sometimes referred as transitional poor households or *middle class* population.
- Solvent class: A solvent household is usually the household with a total monthly income of BDT 19,000 or above. This group also referred as *high income households*.

### 2.10 Data collection procedure

This cross-sectional observational study was carried out on 100 diabetic patients in the Department of Medicine, Shaheed Suhrawardy Medical College Hospital, Dhaka after acceptance of protocol. A total of 100 samples were selected. Subjects or their relatives were briefed about the objectives of the study, risks and benefits, freedom to participate in the study, and confidentiality. Informed consent was obtained accordingly. The present and past history of each case record was evaluated in detail regarding their general & clinical information. The study protocol included a thorough history taking, a sociodemographic profile and complaints related to any complications were noted in detail.

Every consecutive patient who was admitted with fever, cough, and sputum production was evaluated for the presence of CAP. Diagnosis of CAP was made on the basis of history, clinical examination, routine blood parameters (complete blood count including Erythrocyte Sedimentation Rate), chest radiograph and sputum examination. On admission, sputum samples were collected as per standard recommended protocols before the patients received the first dose of antibiotics. In those patients who were unable to expectorate a satisfactory sputum specimen, sputum induction methods were followed. Sputum samples were sent for Gram staining and culture and sensitivity to antibiotics. In addition, sputum was stained by Ziehl-Neelsen staining for the presence of Acid Fast Bacilli (AFB). Patients with CAP but without any bacterial growth in sputum, patients with the growth of fungus or presence of AFB, patients who had already received antibiotics before sputum could be sent for culture sensitivity, aspiration pneumonia, nosocomial pneumonia, and patients on immune-suppressive therapy were excluded from the study.

The pre-structured Case Record Form (CRF) was filled up by the study physician himself. The case definitions of operational variables were described. Patient data such as age, sex, clinical presentation, etc were noted. This questionnaire was used for the collection of information by interviewing patients. All the data collected were checked very carefully to identify errors in collecting data. Data processing work consisted of registration of schedules, editing, coding and computerization, preparation of dummy tables, analysis and matching data. The technical matters of editing, encoding and computerization were looked by researcher.

### 2.11 Variables

#### Independent variables

- Demographic characteristics like age, gender, residence, occupation, economic status
- Clinical manifestation
- Glycemic status
- Frequency of community-acquired pneumonia

Dependent variables:

- Sputum culture & antimicrobial sensitivity pattern

### 2.12 Data analysis

Keeping the research topic in concern, a preset questionnaire was set for data collection. Data for socio-demographic and clinical variables were obtained from all participants by the use of a pre-designed and easily understandable questionnaire. After the collection of all information, these data were checked, verified for consistency and edited for finalized results. Data processing work will consist of registration schedules, editing computerization, preparation of dummy tables, and analyzing and matching the data. After editing and coding, the coded data was directly entered into the computer by using SPSS version 6. Data cleaning validation and analysis were performed using the SPSS/PC software and graph and chart by MS Excel. The result was presented in tables in proportion. A “P” value <0.05 was considered as significant.

#### Methodology Proper

1) This study was done in all medicine wards in ShSMCH
2) Pretesting of questionnaire
3) Finalization of questionnaire
4) Consecutive sampling
5) Consent taking
6) Detailed history
7) Physical examination
8) Investigation
9) Filling the questionnaire by data of collected from patients

#### Flow chart showing the sequence of tasks

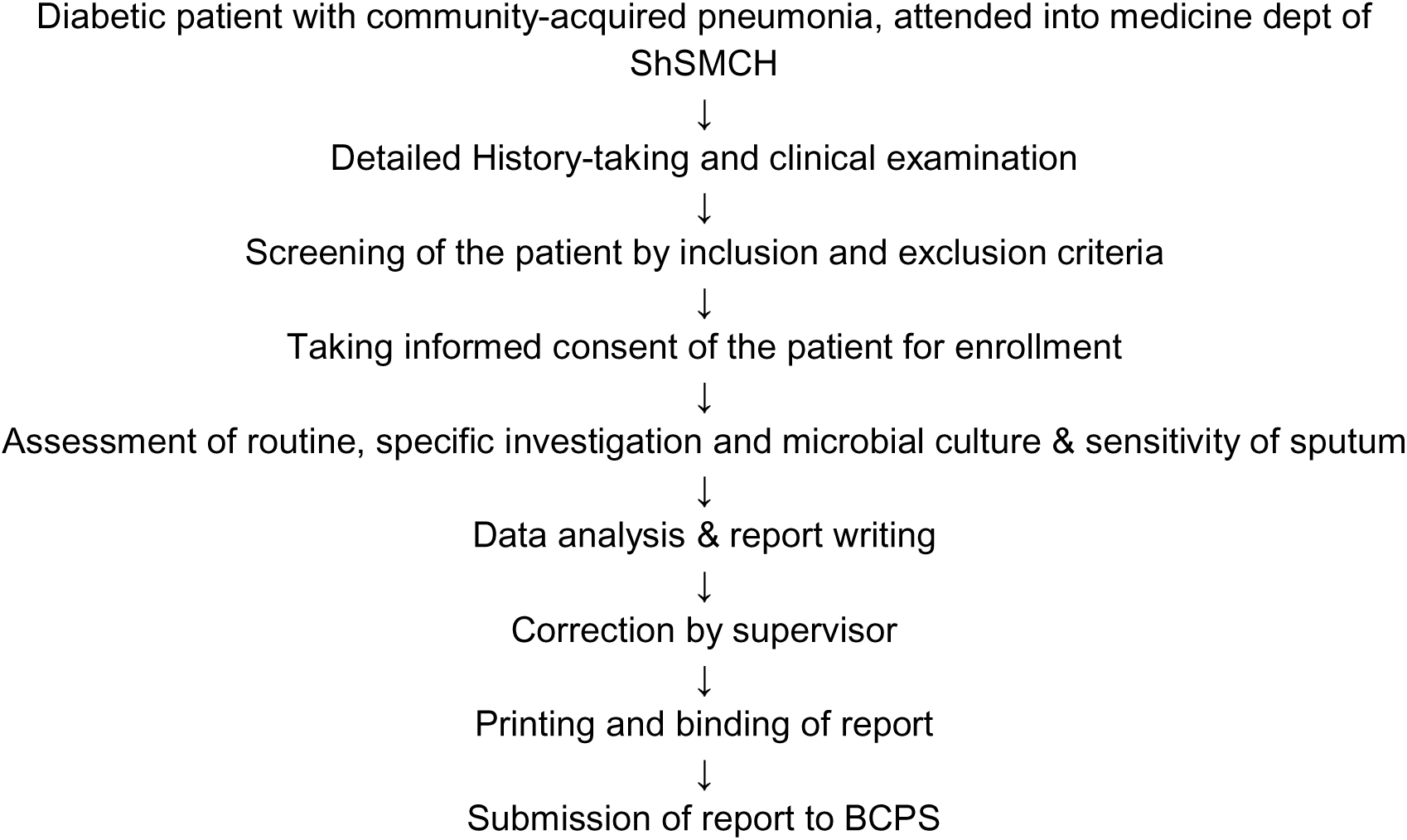

## Results

**Table-1:** Age distribution of the patients (n=100)

| Age (years) | Number of patients | Percentage (%) | Mean ± SD |
| --- | --- | --- | --- |
| 30-40 | 2 | 2.0 |  |
| 41-50 | 30 | 30.0 |  |
| 51-60 | 54 | 54.0 | 49.7 (±9.23) |
| 61-70 | 14 | 14.0 |  |
| **Total** | **100** | **100.0** |  |

In this series, the maximum numbers of patients (54%) were between 51-60 years of age group, next 30.0% were between the age group of 41-50 years, with mean value 49.7 ( ± 9.23) years.

**Table 2:** Gender distribution of the patients (n=100)

| Sex | Frequency | Percentage | M: F |
| --- | --- | --- | --- |
| Male | 62 | 62.0 | 1.6: 1 |
| Female | 38 | 38.0 |  |
| **Total** | **100** | **100.0** |  |

Out of 100 cases 62% were male and 38% were female. Male and female ratio was 1.6:1.

**Figure-1:**
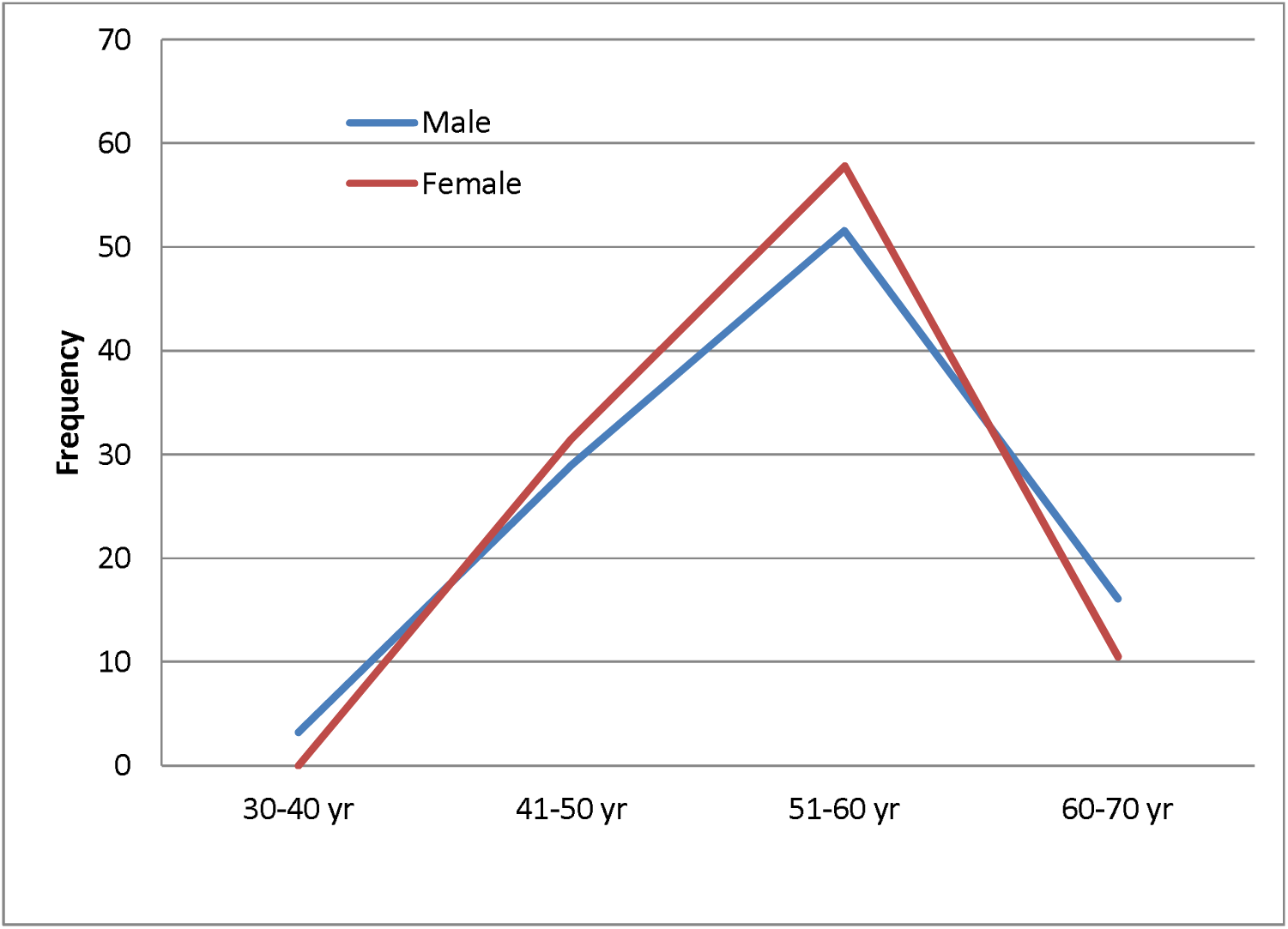
Relationship of frequency of CAP with overall demography (n=100) It shows the frequency of frequency of CAP gradually increased with raised age. The highest incidence of disease was 51-60 years in both male and female.

**Table-3:**
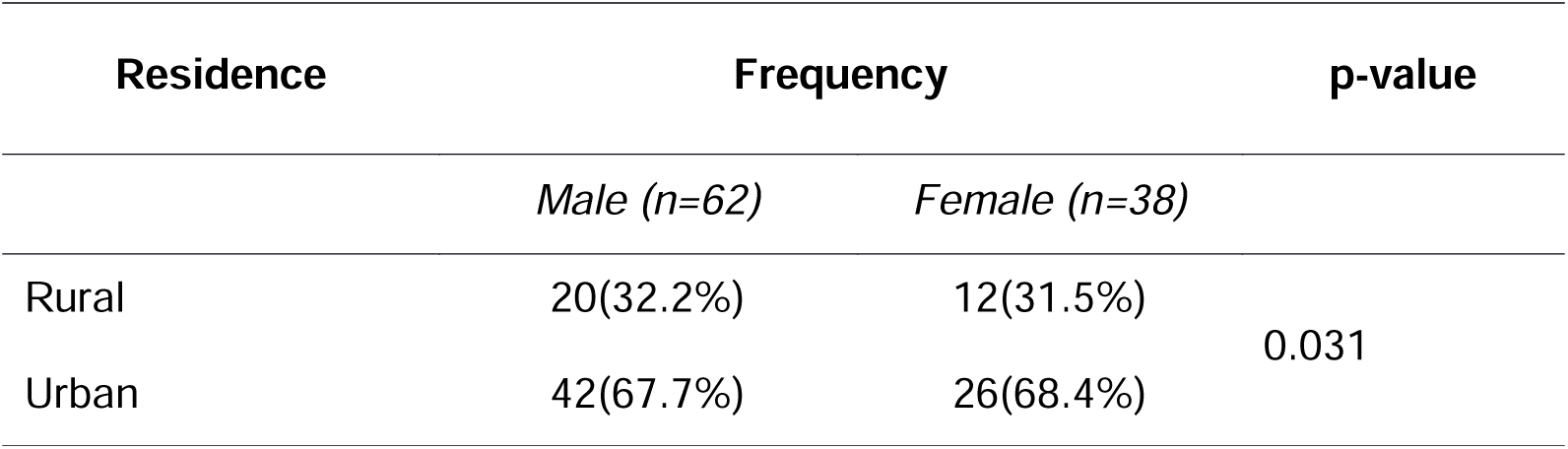
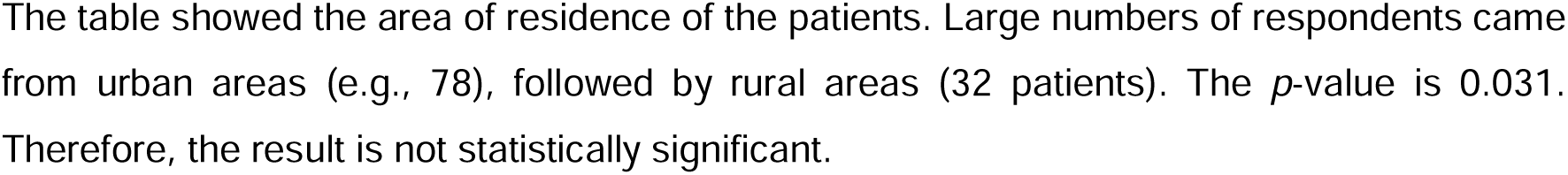
Distribution of patients according to residence (n=100)

**Table 4:** Distribution of the patients according to occupation category (n=100)

| Occupation | Number of patients | Percentage (%) |
| --- | --- | --- |
| Service holder | 13 | 13.0 |
| Businessman | 24 | 24.0 |
| Worker (day laborer) | 25 | 25.0 |
| Housewife | 23 | 23.0 |
| Student | 15 | 15.0 |
| **Total** | **100** | **100.0** |

Many of the patients comprised of daily worker (25.0%) followed by businessman (24.0%), housewife (23.0%) and student 15.0%.

**Figure-2:**
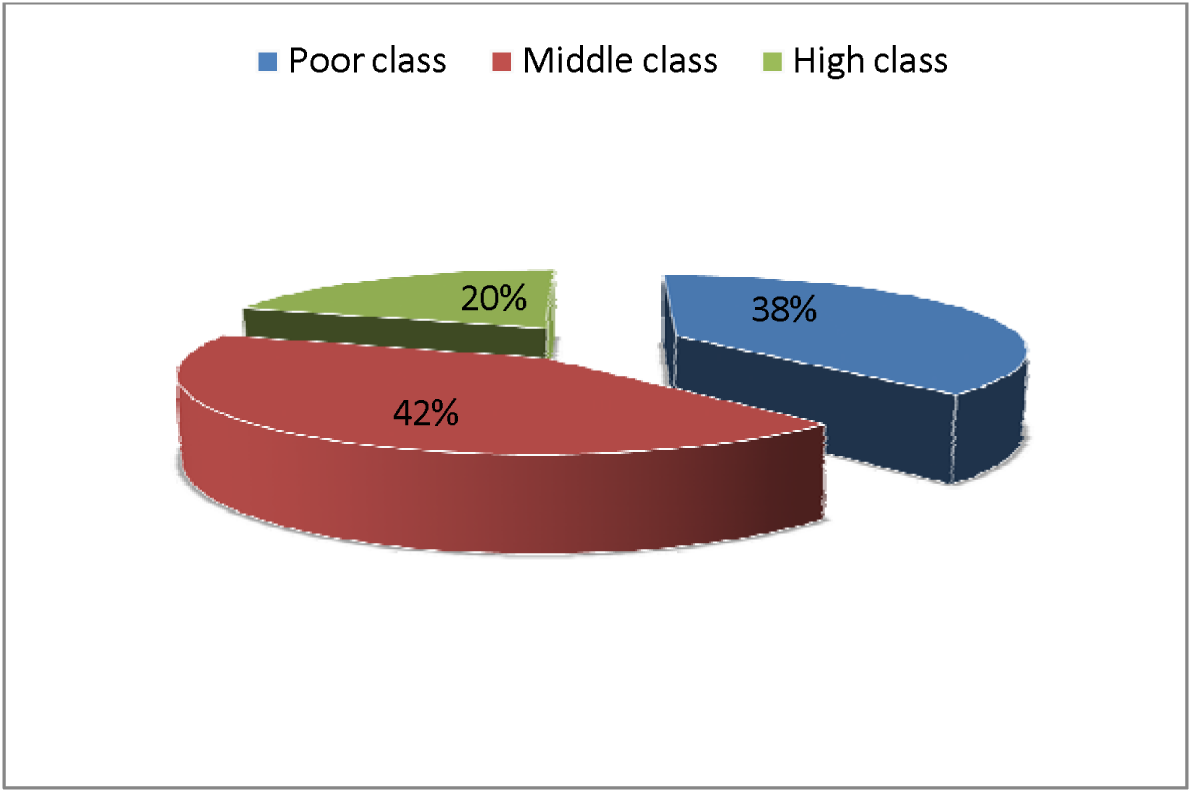
Socioeconomic status of the study population (n=100) Patients are divided into three groups according to operational definition. Among the patients the middle class (42%) comprises the major percentage of the patients, which is followed by poor class (38%) and remaining were in the upper class (20%).

**Table 3.5:** Clinical manifestation of respondents.

| Clinical manifestation | Frequency | Percentage (%) |
| --- | --- | --- |
| High-grade fever, accompanied by sweating | 72 | 72.0 |
| Coughing with sputum | 65 | 65.0 |
| Respiratory distress | 58 | 58.0 |
| Lethargy | 79 | 79.0 |
| Polyuria, fatigue | 65 | 65.0 |
| Chest pain | 46 | 46.0 |
| Acute confusional state | 35 | 35.0 |
| Nausea, vomiting | 46 | 46.0 |
| Loss of appetite | 57 | 57.0 |
| Asymptomatic | 23 | 23. |

**Table 3.4:**
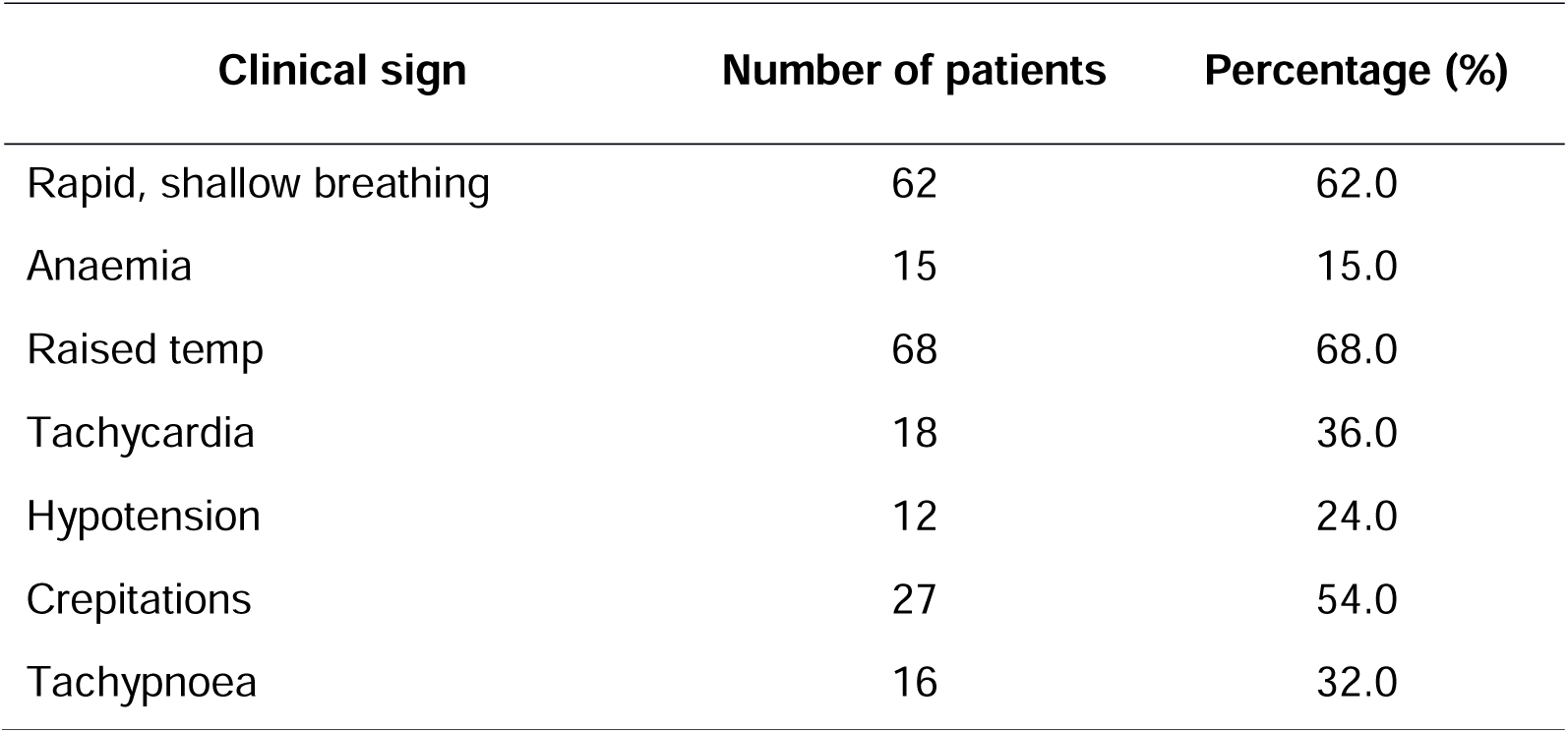
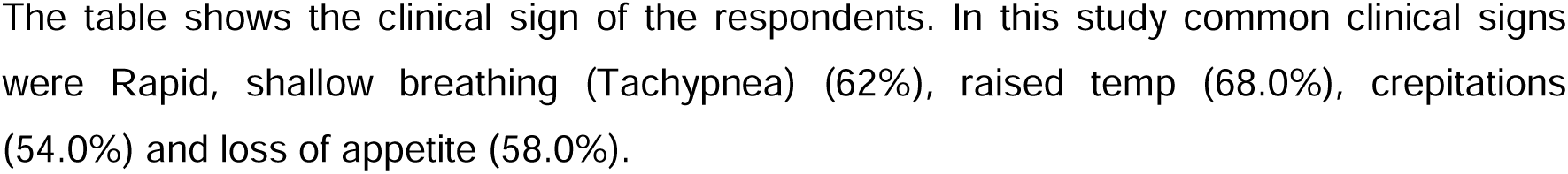
Clinical signs of the respondents.

**Figure 3:**
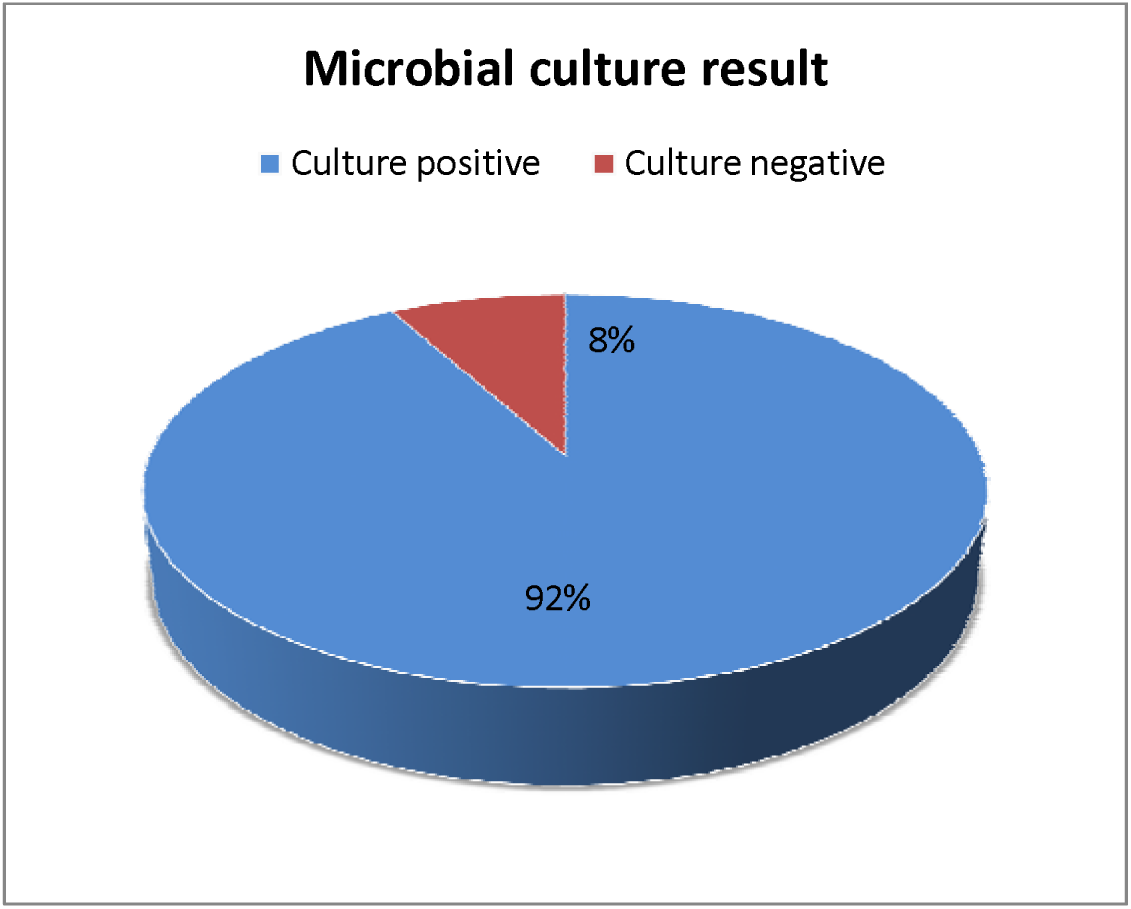
Microbial culture result of community-acquired pneumonia (n=100) A sputum sample was studied to detect the bacterial etiology of pneumonia. Sputum containing more than 25 polymorphonuclear cells and less than 10 epithelial cells per low-power field was subjected to Gram staining and culture. The samples were processed according to standard microbiological practices. Of the 100 sputum samples, 92.0% yielded growth. Among the growths, 34.0% were Gram-positive cocci, 44.0% were Gram-negative bacilli and 14.0% were Gram-negative coccobacilli.

**Table-3.6:**
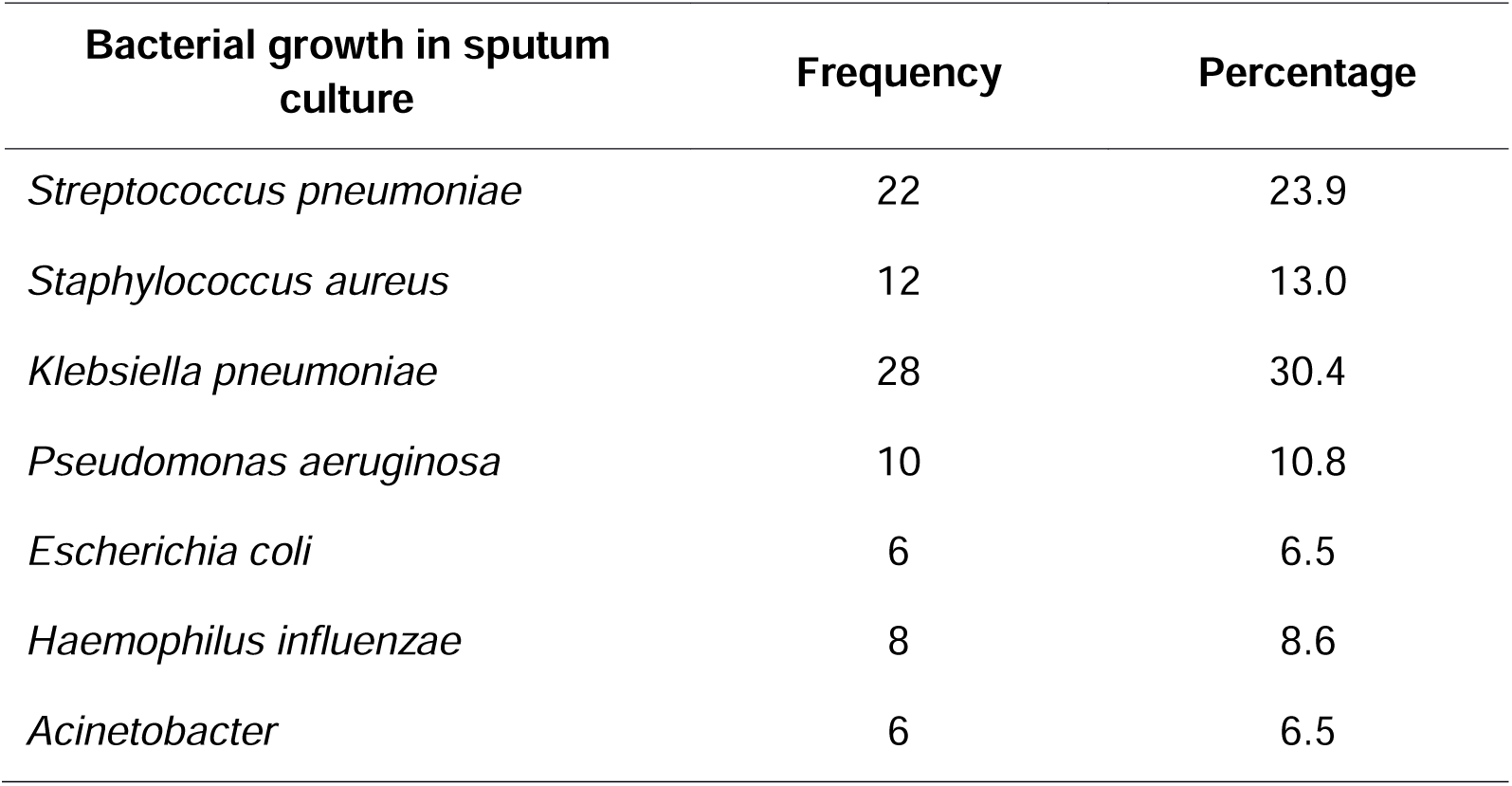
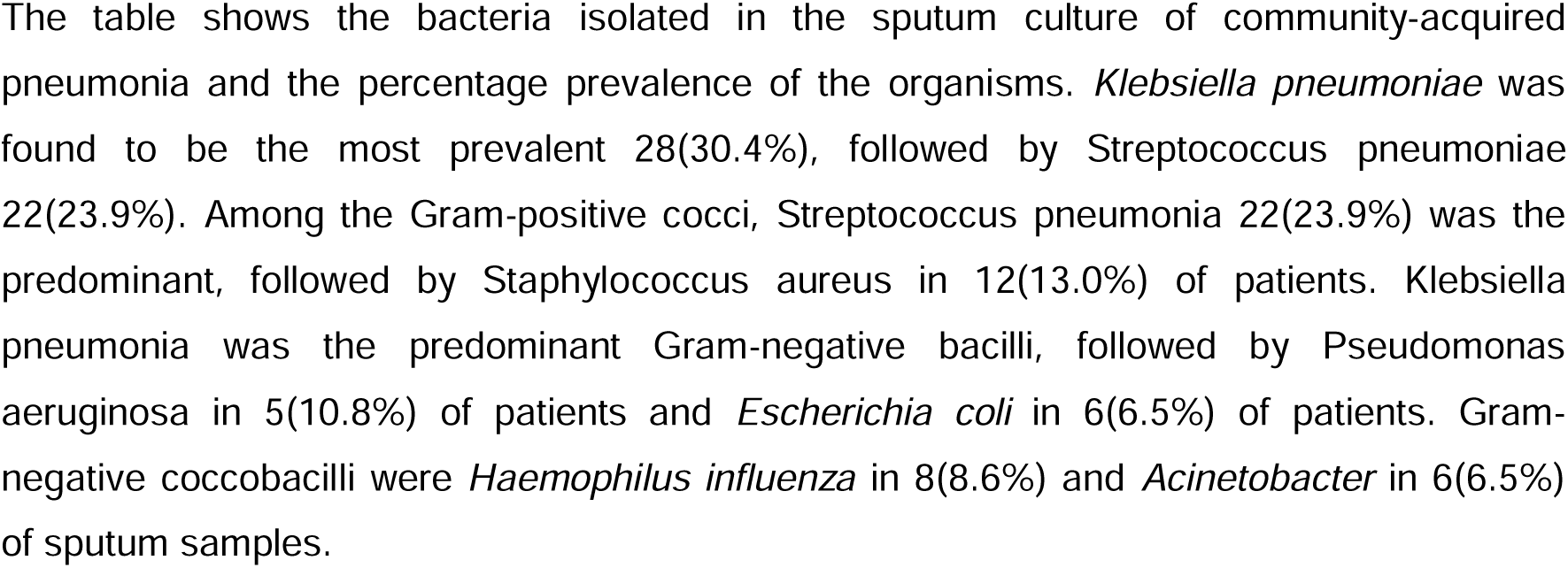
Isolated microorganism from Sputum culture with frequency (n=92)

**Table-3.7:** Sensitivity of common bacterial growth in sputum to different antibiotics (Ceftriaxone, Ceftazidime, Cefixime, Amoxycillin) (n=92)

| Bacterial growth in sputum culture | Sensitivity of Antibiotics (Ceftriaxone, Ceftazidime, Cefixime, Amoxycillin) |  |  |  |
| --- | --- | --- | --- | --- |
|  | Ceftriaxone | Ceftazidime | Cefixime | Amoxycillin |
| *Streptococcus pneumoniae* (n=22) | 10(45.5%) | 12(54.5%) | 8(36.3%) | 0 |
| *Staphylococcus aureus* (n=12) | 4(33.3%) | 6(50.0%) | 6(50.0%) | 2(16.7%) |
| *Klebsiella pneumoniae* (n=28) | 16(57.1%) | 4(14.2%) | 8(28.5%) | 6(21.4%) |
| *Pseudomonas aeruginosa* (n=10) | 2(20.0%) | 0 | 2(20.0%) | 0 |
| *Escherichia coli* (n=6) | 2(33.3%) | 0 | 0 | 0 |
| *Haemophilus influenza* (n=8) | 2(25.0%) | 0 | 2(25.0%) | 0 |
| *Acinetobacter* (n=6) | 0 | 0 | 0 | 2(33.3%) |

**Table-3.8:** Sensitivity of common bacterial growth in sputum to different antibiotics (Amikacin, Ciprofloxacin, Clarithromycin, Imipenem) (n=92)

| Bacterial growth in sputum culture | Sensitivity of Antibiotics (Amikacin, Ciprofloxacin, Clarithromycin, Imipenem) |  |  |  |
| --- | --- | --- | --- | --- |
|  | Amikacin | Ciprofloxacin | Clarithromycin | Imipenem |
| *Streptococcus pneumonia* (n=22) | 4(18.1%) | 4(18.1%) | 10(45.5%) | 18(81.8%) |
| *Staphylococcus aureus* (n=12) | 0 | 0 | 8(66.7%) | 12(100.0%) |
| *Klebsiella pneumonia* (n=28) | 10(35.7%) | 4(14.2%) | 8(28.5%) | 24(85.7%) |
| *Pseudomonas aeruginosa* (n=10) | 8(80.0%) | 2(20.0%) | 0 | 8(80.0%) |
| *Escherichia coli* (n=6) | 4(66.7%) | 0 | 2(33.3%) | 6(100.0%) |
| *Haemophilus influenza* (n=8) | 6(75.0%) | 0 | 2(25.0%) | 6(75.0%) |
| *Acinetobacter* (n=6) | 6(100.0%) | 2(33.3%) | 0 | 6(100.0%) |

Bacterial antibiotic sensitivity pattern to Amikacin, Ciprofloxacin, Clarithromycin and Imipenem were as follows- *Klebsiella pneumonia* (35.7%, 14.2%, 28.5%, 85.7% respectively), *Streptococcus pneumonia* (18.1%, 18.1%, 45.5%, 100% respectively), *Staphylococcus aureus* (0%, 0%, 66.7%, 100% respectively), *Pseudomonas* species (80.0%, 20.0%, 0%, 80.0% respectively), *Escherichia coli* (66.7%, 0%, 33.3%, 100.0% respectively), *Haemophilus influenza* (75.0%, 0%, 25.0%, 75.0% respectively), *Acinetobacter* (100%, 33.3%, 0%, 100% respectively).

**Figure-4:**
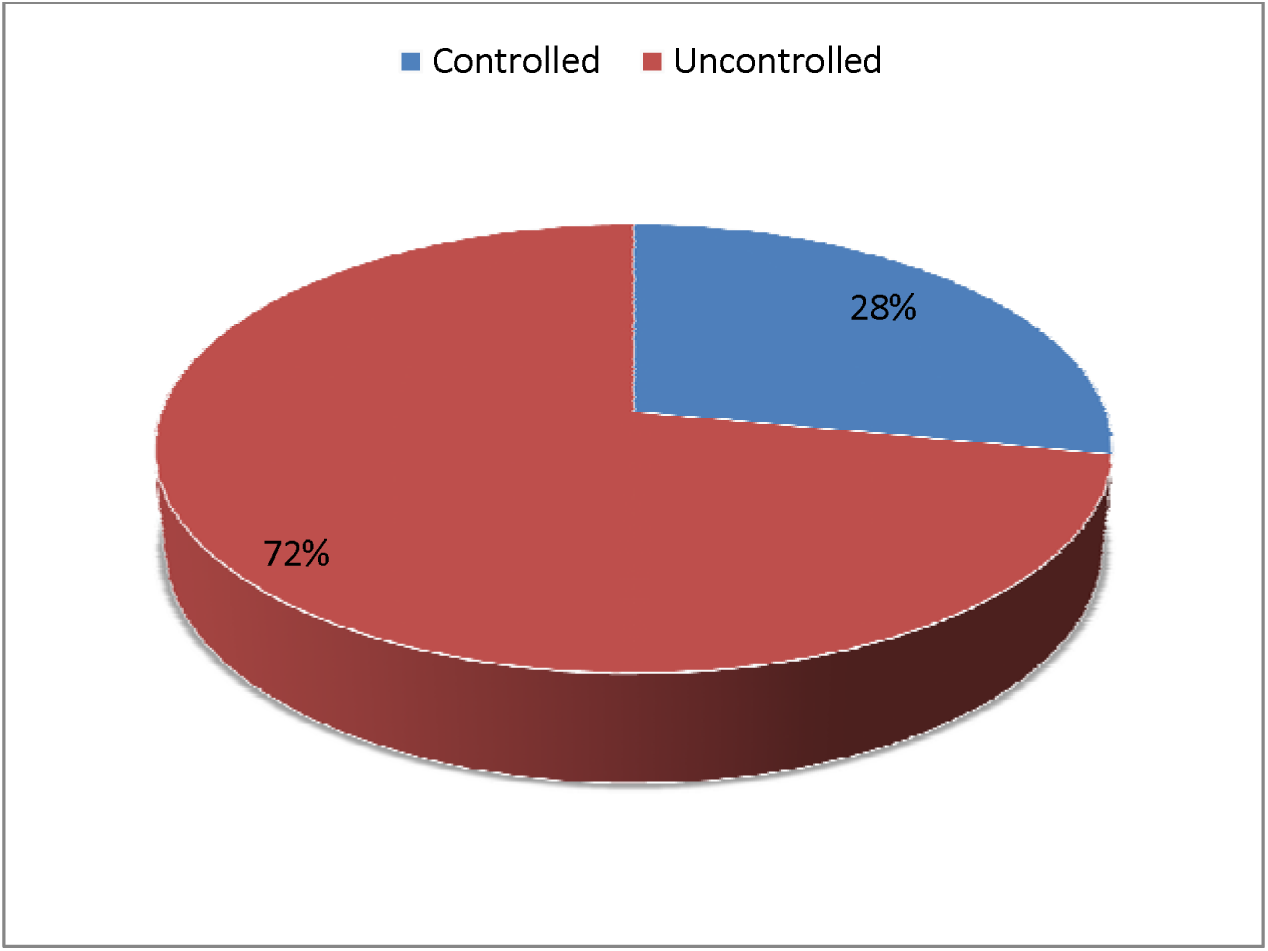
Status of DM (glycemic status) during hospitalization (n=100) At the time of hospitalization all available investigation reports, previous medical records, and the status of DM (glycemic status) were evaluated meticulously. The present study shows that the maximum patients (72.0%) had uncontrolled glycemic status.

**Table-3.9:** Assessment of HbA1c (%) levels in study population (n=100)

| HbA1c (%) | Frequency |  | Total | p-value |
| --- | --- | --- | --- | --- |
|  | Male (n=62) | Female (n=38) |  |  |
| 5-6% | 8(12.9%) | 5(13.1%) | 13 | 0.032 |
| 6-7% | 10(16.1%) | 5(13.1%) | 15 |  |
| 7-8% | 16(25.8%) | 12(31.5%) | 28 |  |
| 8-9% | 28(45.1%) | 16(42.1%) | 44 |  |

**Table- 3.9: Assessment of HbA1c (%) levels in study population (n=100)**
| HbA1c (%) | Frequency |  | Total | p-value |
| --- | --- | --- | --- | --- |
|  | Male (n=62) | Female (n=38) |  |  |
| <7% | 18(29.0%) | 10(26.3%) | (28.0%) | 0.032 |
| >7% | 44(70.9%) | 28(73.6%) | (72.0%) |  |

Regarding the operational definition of glycemic control, less than 7% regarded as good, >7% is poor status of glycemic control. In this study, the majority of patients 72.0% had >7% of HbA1c (%) levels during the study period, and 28.0% of patients recorded <7%. The result is not significant at $p < .05$ . In this series we also found that glycemic control was comparatively good in male than female subjects (29.0% vs. 26.3%). The p-value is 0.03271.

**Table 3.10:** Correlation of glycaemic status with bacterial growth in sputum of the diabetic patients (n=100)

| Bacterial growth in sputum culture | Glycemic status (HbA1c level) |  | Total |
| --- | --- | --- | --- |
|  | <7% | >7% |  |
| *Streptococcus pneumonia* (n=22) | 6(27.2%) | 16(72.8%) | 22 |
| *Staphylococcus aureus* (n=12) | 6(50.0%) | 6(50.0%) | 12 |
| *Klebsiella pneumonia* (n=28) | 10(35.7%) | 18(64.2%) | 28 |
| *Pseudomonas aeruginosa* (n=10) | 4(40.0%) | 6(60.0%) | 10 |
| *Escherichia coli* (n=6) | 0 | 6(100.0%) | 6 |
| *Haemophilus influenza* (n=8) | 2(25.0%) | 6(75.0%) | 8 |
| *Acinetobacter* (n=6) | 0 | 6(100.0%) | 6 |
| **Total** | **28** | **72** |  |

In this study, most of the growth of all the bacteria (*Klebsiella* 64.2%, *Staphylococcus* 72.8%, *Pseudomonas* 60.0%, *Acinetobacter* 100%, *E. coli* 100%) occurred in patients with poor glycemic control (HbA1c $\geq 7.0\%$ ).

**Figure 5:**
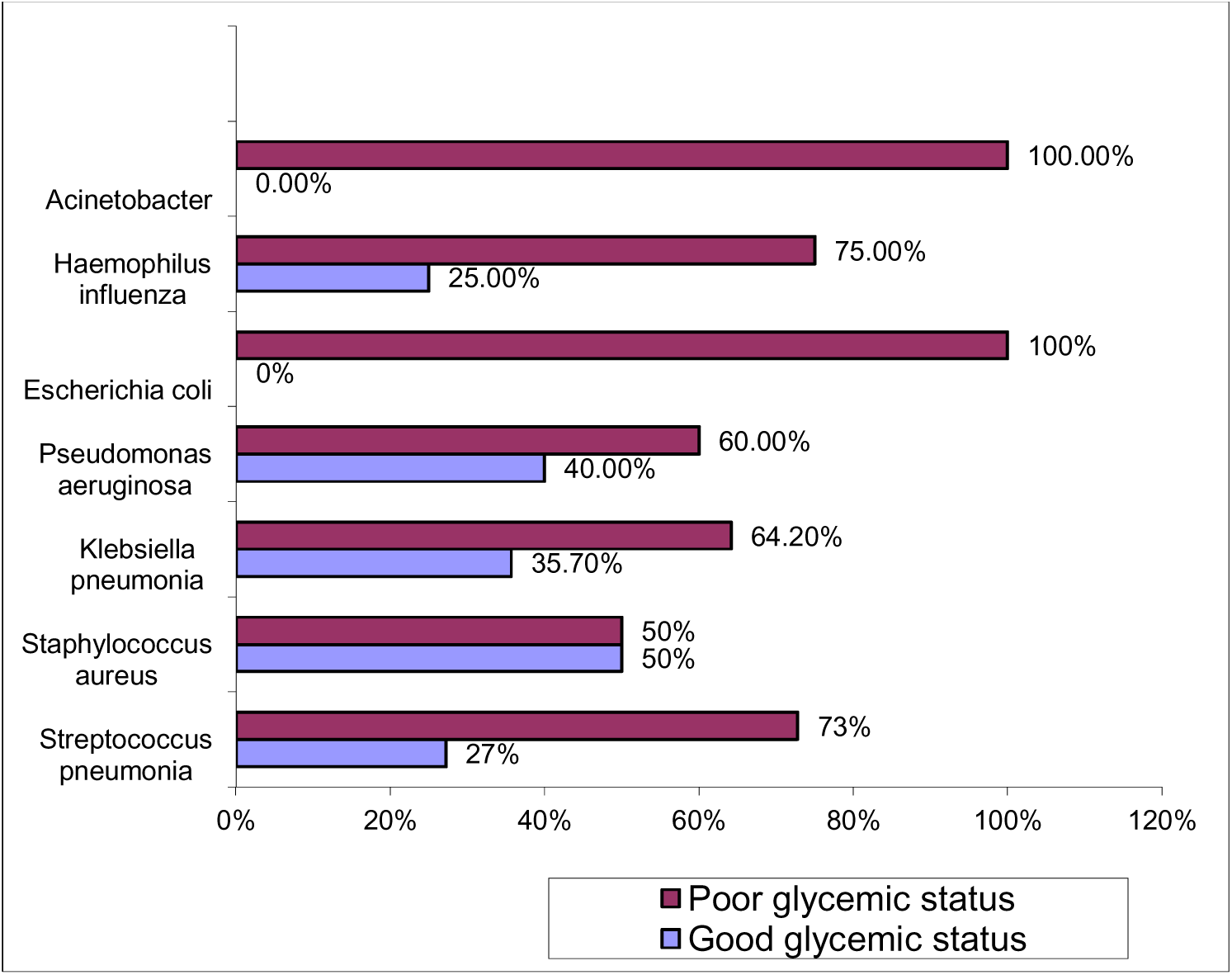
Spectrum of bacterial growth in relation to glycemic status of diabetes patients (n=100) It shows the Spectrum of bacterial growth in relation to the glycemic status of diabetes patients. Diabetes is a disease, which has profound deleterious effects on immunity. As a result, it predisposes to a variety of infections. Regarding glycemic status, most of the bacterial growth was isolated in patients with uncontrolled DM as evidenced by HbA1c ≥7.0%. This is because uncontrolled DM causes immunosuppression leading to an increased chance of any infection including pneumonia.

## Discussion

This cross-sectional study was conducted in Shaheed Suhrawardy Medical College Hospital over six months to see the microorganisms most commonly causing community-acquired pneumonia in diabetic patients. In this study, the maximum number of patients (54%) was between 51-60 years of age group, with a mean value of 49.7 (±9.23) years. Out of 100 cases, 62% were male and 38% were male. Male and female ratio was 1.6:1. Large numbers of respondents came from urban areas (e.g., 78), followed by rural areas (32 patients).

All this finding is like some other studies. In a study, the mean age (±SD) of the patients was 56.3(±12.2)^21^. In another study, the average age in SG was 57.93±9.71 yrs^8^. In this study, the male and female ratio was 1.6:1. This finding is similar to some other studies where 62% of DM patients with CAP were male^23^ and in another study, 67.0% were male^12^. This may be because male patients may have more incidence of CAP and have more access to health facilities than females. The mean age of the patients in this study was around 55 years which was lower than other studies in the Western world where the mean age of the DM patients with CAP was around 72 years, which may be due to the difference in average life span in different countries^23^.

The BTS Guideline for Diagnosing CAP is as Follows: a) symptoms of an acute lower respiratory tract illness (cough with or without expectoration, shortness of breath, pleuritic chest pain) for less than 1 week; and b) At least one systemic feature (temperature >37.7°C, chills, and rigors, and/or severe malaise); and c) New focal chest signs on examination (bronchial breath sounds and/or crackles) with d) No other explanation for the illness^14^. The present study gives the impression that High-grade fever, accompanied by sweating, cough, lethargy, and respiratory distress, was the commonest presentation in patients of DM (72%, 65%, 79%, and 58% of patients respectively) as shown in the table. It was evident that 23% of patients were asymptomatic. Findings consistent with the result of other studies. In a study fever as a presenting symptom was absent in 10.7% of the patients. Cough was present in 96.4%, 89.3% had a history of sputum production,57% had breathlessness at presentation and 53.6% had a history of chest pain^14^.

In this study of the 100 sputum samples, 92.0% yielded growth. Among the growths, 34.0% were Gram-positive cocci, 44.0% were Gram-negative bacilli and 14.0% were Gram-negative coccobacilli. *Klebsiella pneumoniae* was found to be the most prevalent 28(30.4%), followed by Streptococcus pneumoniae 22(23.9%). Among the Gram-positive cocci, Streptococcus pneumonia 22(23.9%) was the predominant, followed by Staphylococcus aureus in 12(13.0%) of patients.

This finding is similar to other studies conducted in Bangladesh^12, 24, 25^ but somehow different from another study in India where the majority of growth was Pseudomonas followed by Staphylococcus aureus^26^. It has been suggested that patients with DM have an increased rate of colonization and adherence of gram-negative bacteria to the upper respiratory epithelium. Their aspiration of these bacteria to the lung may be facilitated by the use of anti-ulcerants and diabetic gastroparesis^27, 28^. Diabetic patients are also at increased risk of staphylococcal pneumonia as because the rate of nasal carriage of *Staphylococcus* in diabetic patients is 30% compared to 11% in non-diabetic individuals^29^. Similarly, another study in Bangladesh reported that *Klebsiella pneumonia* was the most commonly isolated organism from sputum samples. It is followed by *Streptococcus pneumoniae*, *Staphylococcus aureus, E. Coli* and *Pseudomonas aeruginosa*^21^.

In this study bacterial antibiotic sensitivity patterns to ceftriaxone, Ceftazidime, Cefixime, and Amoxycillin were as follows-*Klebsiella pneumonia* (57.1%, 14.2%, 28.5%, 21.4% respectively), *Streptococcus pneumonia* (45.5%, 54.5%, 36.3%, 0% respectively), *Staphylococcus aureus* (33.3%, 50.0%, 50.0%, 16.7% respectively), *Pseudomonas* species (20.0%, 0%, 20.0%, 0% respectively), *Escherichia coli* (33.3%, 0%, 0%, 0% respectively), *Haemophilus influenza* (25.0%, 0%, 25.0%, 0% respectively), *Acinetobacter* (0%, 0%, 0%, 33.3% respectively).

Findings accordance with the result of other studies in Bangladesh, e.g., bacterial antibiotic sensitivity pattern to ceftriaxone, ciprofloxacin, amikacin and imipenem were as follows: *Klebsiella pneumoniae* (19%, 47%, 74%, 96% respectively), *Staphylococcus aureus* (1 1%, 33%, 78%, 67% respectively), *Pseudomonas* species(19%, 75%, 81%, 88% respectively), *Acinetobacter* (0%, 0%, 20%, 50% respectively), *Escherichia coli* (22%, 22%, 100%, 100% respectively). Most of the bacterial growth in the sputum of all the bacteria was found in patients with HbA1c ≥7.0%^12^.

It is worth noting that the growth of relatively rare organisms like *Acinetobacter* was quite high in diabetic patients. Moreover, growth of St*reptococcus pneumoniae* is negligible compared to conventional findings in nondiabetic patients^30^. Antibiotic sensitivity pattern showed that most of the bacteria including almost 80% of *Klebsiella* were resistant to ceftriaxone. This is similar to other studies in Bangladesh^24^ and India^30^ but different from another study finding where almost 90% of the *Klebsiella* was sensitive to ceftriaxone^25^.

A similar study in Bangladesh reported regarding the antimicrobial sensitivity pattern of isolated organisms, all the isolates (100%) of *Klebsiella pneumoniae* from diabetic patients with CAP were resistant to Co-amoxiclav, 66.7% to Levofloxacin, 55.6% to Clarithromycin and 11.1% to Ceftriaxone and Ceftazidime. All isolates of *Staphylococcus aureus* from diabetic patients with CAP were sensitive to Ceftriaxone, Imipenem, Meropenem and 50% sensitive to Ceftazidime, Clarithromycin and Levofloxacin and all were resistant to Coamoxiclav. *E. coli* was isolated from only diabetic patients. All *E. coli* isolates were sensitive to Ceftazidime, Imipenem, and Meropenem, 50% to Ceftriaxone and Levofloxacin and all were resistant to Co-amoxiclav. All the *Pseudomonas aeruginosa* isolates were from diabetic patients with CAP, which were sensitive to Ceftazidime, Imipenem, and Meropenem and were resistant to Co-amoxiclav, Ceftriaxone, Clarithromycin, and Levofloxacin^21^.

At the time of hospitalization all available investigation reports, previous medical records, status of DM (glycemic status) were evaluated meticulously. The present study shows that the maximum patients (72.0%) had uncontrolled glycemic status. In this study, most of the growth of all the bacteria (*Klebsiella* 64.2%, *Staphylococcus* 72.8%, *Pseudomonas* 60.0%, *Acinetobacter* 100%, *E. coli* 100%) occurred in patients with poor glycemic control (HbA1c ≥7.0%).

Findings per another study regarding glycemic status, most of the bacterial growth was isolated in patients with uncontrolled DM as evidenced by HbA1c ≥ 7.0%. This is because uncontrolled DM causes immunosuppression leading to an increased chance of any infection including pneumonia^12^. Hyperglycemia impairs a wide range of functions in neutrophils and macrophages, therefore uncontrolled diabetes is associated with increased susceptibility to various infections like pneumonia. So, whenever possible, treatment of CAP should be guided by sputum culture and sensitivity tests, and for empirical treatment of CAP in diabetic patients, rationale antibiotics should be considered.

## Conclusions

Sputum culture is an essential step in knowing the microbiological profile and drug sensitivity among patients with CAP. This study’s results suggest that CAP in diabetic patients is more frequent due to gram-negative bacilli like *Klebsiella pneumoniae*, *Pseudomonas* species, and also *Staphylococcus aureus,* and mostly they are resistant to commonly used antibiotics like ceftriaxone and ciprofloxacin. According to the results, this study emphasizes the need for strict glycemic control to prevent any infection in diabetic patients. Community-acquired pneumonia (CAP) is a common and potentially serious illness. It is associated with considerable morbidity and mortality, particularly in older patients and those with poor glycemic control. The spectrum of potential pathogens known to cause pulmonary infections in immunocompromised individuals has grown as a result of intensified immunosuppression, prolonged patient survival, the emergence of antimicrobial-resistant pathogens, and improved diagnostic assays. Survival has improved with the availability of newer antimicrobial agents. Pneumonia in diabetic patients is often atypical, caused by more virulent organisms, and associated with increased antibiotic resistance. Though various studies have assessed the common microbial agents, and susceptibility patterns in Community-acquired Pneumonia (CAP), most of them were from the western world. Studies in this regard are also lacking in our country.

## Limitations of the study

1) Small sample size: only patients admitted in the Department of Medicine, ShMCH were taken for the study. So, this will not reflect the overall picture of the country. A large-scale study needs to be conducted to reach a definitive conclusion.
2) Acute signs and symptoms were limited by the fact that patients were evaluated on admission or during hospitalization; it is difficult to generalize the findings to the reference population.
3) The majority of the patients were poor and ignorant, many had no proper check-up. Patients come to tertiary centres after the development of some hazards, and complications. So, some hazards and risk factors are subsistent before and after treatment.
4) Samples were taken by a purposive method in which questions of personal biases might arise.

### Recommendation

- All patients with diabetes or caregivers should understand their condition and the resources available to optimize their general health, diabetes management, and general care.
- Education is essential as an empowerment strategy for self-management of diabetes and prevention or reduction of complications. Education is based on identified individual needs, risk factors, ulcer status, available resources and ability to heal
- Health care professionals should assess all patients with diabetic related complications for signs and symptoms of infection and facilitate appropriate diagnostic testing and treatment at primary, secondary level health care centre.

### Ethical Considerations and Quality Measures

This study was conducted in accordance with the ethical principles outlined in the Declaration of Helsinki. Ethical approval was obtained from the Ethics Committee of Shaheed Suhrawardy Medical College Hospital, Dhaka, Bangladesh before commencing the study. All participants provided informed written consent prior to their inclusion in the study. The purpose of the research, the procedures involved, and the potential risks and benefits were explained to all participants. Confidentiality of patient information was maintained, and data were anonymized to protect the privacy of the participants. Participation was voluntary, and patients had the right to withdraw from the study at any point without any repercussions.

In order to ensure proper quality of the data collection procedure, firstly a work manual was made. Then a sample size was selected. A standard questionnaire was made. Then the questionnaire was pretested. The pretest ensures that the respondents are able to understand the questionnaire and answer accordingly.

## Data Availability

All data produced in the present study are available upon reasonable request to the authors

## Acknowledgements

The authors express their heartfelt gratitude to the Department of Medicine, Shaheed Suhrawardy Medical College Hospital, Dhaka, Bangladesh for providing the necessary facilities and support to conduct this research. Special thanks are extended to the clinical and laboratory staff for their assistance in data collection and laboratory procedures. The authors are also grateful to the patients and their families for their cooperation and willingness to participate in the study. Finally, sincere appreciation goes to colleagues and peers for their valuable feedback and guidance throughout the research process.

## Notes

### Competing Interest Statement

The authors have declared no competing interest.

### Funding Statement

The study did not receive any funding

### Author Declarations

Ethical approval was obtained from the Ethics Committee of Shaheed Suhrawardy Medical College Hospital, Dhaka, Bangladesh before commencing the study.

## References

1. Roglic G. Global report on diabetes. World Health Organization. 2016;1–54.

2. Bommer C, Sagalova V, Heesemann E, Manne-Goehler J, Atun R, Bärnighausen T, et al. Global economic burden of diabetes in adults: projections from 2015 to 2030. Diabetes Care. 2018 May;41(5):963–70.

3. King H, Aubert RE, Herman WH. Global burden of diabetes, 1995-2025: prevalence, numerical estimates, and projections. Diabetes Care. 1998 Sep;21(9):1414–31.

4. Dutt J, Leena A. Study of pneumonia in diabetic patients. Int J Med Sci Public Health. 2014;3(8):974–6.

5. Kahn CR, Weir GC, King GL, Jacobson A, Smith R, Moses A. Joslin’s diabetes mellitus. 14th ed. Ovid Technologies; 2005.

6. Eirini T, Argyriou K, Makris D, Zakynthinos E. Type, course and outcome of community acquired infection in hospitalized diabetes. Microbiol Res. 2010;1(1):1–5.

7. Kornum JB, Thomsen RW, Riis A, Lervang HH, Schønheyder HC, Sørensen HT. Type 2 diabetes and pneumonia outcomes: a population-based cohort study. Diabetes Care. 2007;30:2251–7.

8. Bhambar S, Deore P, Rathod R, Janrao S. Pneumonia in diabetics: clinico-bacteriological profile and outcome. Int J Med Health Res. 2017;3(6):62–6.

9. Peleg AY, Weerarathna T, McCarthy JS, Davis TM. Common infections in diabetes: pathogenesis, management and relationship to glycaemic control. Diabetes Metab Res Rev. 2007 Jan;23(1):3–13.

10. Fine MJ, Smith MA, Carson CA, Mutha SS, Sankey SS, Weissfeld LA, et al. Prognosis and outcomes of patients with community-acquired pneumonia: a meta-analysis. JAMA. 1995;274:134–40.

11. Ljubić S, Metelko Z, Car N, Roglić G, Drazić Z. Reduction of diffusion capacity for carbon monoxide in diabetic patients. Chest. 1998;114:1033–5.

12. Ahmed JU, Hossain MD, Rahim MA, Afroz F, Musa AKM. Bacterial etiology and antibiotic sensitivity pattern of community acquired pneumonia in diabetic patients: experience in a tertiary care hospital in Bangladesh. BIRDEM Med J. 2017;7(2):101–5.

13. Endeman H. Clinical characteristics and innate immunity in patients with community-acquired pneumonia. [Thesis]. Utrecht: University of Utrecht; 2009. p. 7–36.

14. Niyas VKM, Kumar KGS. Community acquired pneumonia in type 2 diabetes mellitus: a study of clinical and bacteriological profile. J Evid Based Med Healthc. 2016;3(17):656–61.

15. Ahasan H, Islam M, Alam M, Miah M, Nur Z, Mohammed F, et al. Prevalence and risk factors of type 2 diabetes mellitus among secretariat employees of Bangladesh. J Med. 2011;12(2):125–30.

16. Rahman MS, Akter S, Abe SK, Islam MR, Mondal MNI, Rahman JAMS, et al. Awareness, treatment, and control of diabetes in Bangladesh: a nationwide population-based study. PLoS One. 2015;10(2):e0118365.

17. Kaysin A, Viera AJ. Community-acquired pneumonia in adults: diagnosis and management. Am Fam Physician. 2016;94(9):698–706.

18. Musher DM, Thorner AR. Community-acquired pneumonia. N Engl J Med. 2014;371:1619–28.

19. Fleming DM, Crombie DL, Cross KW. Disease concurrence in diabetes mellitus: a study of concurrent morbidity over 12 months using diabetes mellitus as an example. J Epidemiol Community Health. 1991;45:73–7.

20. Woodhead MA, Macfarlane JT, McCracken JS, Rose DS, Finch RG. Prospective study of aetiology and outcome of pneumonia in the community. Lancet. 1987;27:671–4.

21. Saibal MAA, Rahman SHZ, Nishat L, Sikder NH, Begum SA, Islam MJ, et al. Community-acquired pneumonia in diabetic and non-diabetic hospitalized patients: presentation, causative pathogens and outcome. Bangladesh Med Res Counc Bull. 2012;38:98–103.

22. Innes JAA, Reid PT. Respiratory diseases. In: Boon NA, Colledge NR, Walker BR, Hunter JAA, editors. Davidson’s Principles & Practice of Medicine. 20th ed. London: Churchill Livingstone; 2006. p. 689.

23. Martins M, Boavida JM, Raposo JF, Froes F, Nunes B, Ribeiro RT, et al. Diabetes hinders community-acquired pneumonia outcomes in hospitalized patients. BMJ Open Diabetes Res Care. 2016;4(1):e000181.

24. Hossain MD, Ahmed JU, Musa AKM. Sputum culture and drug sensitivity pattern of community-acquired pneumonia in diabetic patients and their correlation with glycaemic status. Chest. 2010;138:593A.

25. Saibal MAA, Rahman SHZ, Nishat L, Sikder NH, Begum SA, Islam MJ, et al. Community-acquired pneumonia in diabetic and non-diabetic hospitalized patients: presentation, causative pathogens and outcome. Bangladesh Med Res Counc Bull. 2012;38:98–103.

26. Shah BA, Singh G, Naik MA, Dhobi GN. Bacteriological and clinical profile of community-acquired pneumonia in hospitalized patients. Lung India. 2010;27(2):54–7.

27. Heyland D, Mandell LA. Gastric colonization by gram-negative bacilli and nosocomial pneumonia in the ICU. Chest. 1992;101:187–92.

28. Ljubic S, Balachandran A, Pavlic-Renar I, Barada A, Metelko Z. Pulmonary infections in diabetes mellitus. Diabetologia Croatica. 2004;33(4):115–24.

29. Lipsky BA, Pecoraro RE, Chen MS. Factors affecting staphylococcal colonization among NIDDM outpatients. Diabetes Care. 1987;10:403–9.

30. Acharya V, Padyana M, Unnikrishnan B, Anand R, Acharya P, Juneja DJ. Microbiological profile and drug sensitivity pattern among community-acquired pneumonia patients in tertiary care centre in Mangalore, coastal Karnataka, India. J Clin Diagn Res. 2014;8(6):4–6.

31. Hamilton EJ, Martin N, Makepeace A, Sillars BA, Davis WA, et al. Incidence and predictors of hospitalization for bacterial infection in community-based patients with type 2 diabetes: the Fremantle diabetes study. PLoS One. 2013;8(3):e60502.

